# Intracortical microstimulation in humans: a decade of safety and efficacy

**DOI:** 10.1101/2025.08.11.25332271

**Authors:** Charles M. Greenspon, Taylor G. Hobbs, Ceci Verbaarschot, Ali H. Alamri, Natalya D. Shelchkova, Robin Lienkämper, Joel Ye, Tyler W. Simpson, Jeffrey M. Weiss, David M. Weir, Debbie E. Harrington, Ashley Van Driesche, David Satzer, Giacomo Valle, Lee E. Miller, Nicholas G. Hatsopoulos, Jorge Gonzalez-Martinez, Peter C. Warnke, John E. Downey, Michael L. Boninger, Jennifer L. Collinger, Robert A. Gaunt

**Author notes:** Indicates equal contributions (Charles M. Greenspon, Taylor G. Hobbs).

## Abstract

**Background:** Intracortical microstimulation (ICMS) of somatosensory cortex can restore a sense of touch to people with spinal cord injury (SCI). In this early-feasibility clinical trial, we evaluate the safety, efficacy, and longevity of ICMS as there is a paucity of such long-term studies in humans. This information is crucial to the development of clinical neuromodulation devices, particularly for restoring touch, hearing, and vision.

**Methods:** ICMS was delivered to five participants with SCI who were each implanted with two Blackrock NeuroPort microelectrode arrays in the hand representation of Brodmann’s Area 1. Across implant durations spanning two to ten years, we measured single-electrode detection thresholds, projected fields, quality reports, and electrode health characteristics. ICMS-related adverse events were documented throughout.

**Results:** Over 168 million ICMS pulses were delivered across a combined implant duration of 24 years without any serious adverse events or direct negative effects on electrode health. ICMS consistently evoked sensations localized to the hand. Rarely, sensations persisted for brief periods after stimulation offset (3–25 events across participants). ICMS detection thresholds increased slowly over time (∼3.5 μA/year), but 62±15% of the electrodes still reliably evoke tactile sensations (∼25% decrease in functional electrodes), including 55% of the electrodes after 10 years in one participant. The quality and projected field coverage of ICMS-evoked sensations were both consistent.

**Conclusions:** Delivering ICMS to somatosensory cortex was safe and reliable, consistently evoking informative somatosensory percepts as long as 10 years, demonstrating the clinical promise of ICMS for sensory restoration. (Funded by NIH; ClinicalTrials.gov: NCT01894802).

## Introduction

Spinal cord injury (SCI) leads to a loss of voluntary movement and sensation below the level of injury, resulting in a reduction in independence, quality of life and, often, an absence from the workforce^1^. For decades, people with tetraplegia have described improved arm and hand function as their primary rehabilitation goal^2,3^. Recently, intracortical brain-computer interfaces (iBCIs) have emerged as an approach to restoring motor function but they typically lack the somatosensory feedback that enables our native limb to perform dexterous movements^4^. Intracortical microstimulation (ICMS) of the somatosensory cortex can overcome this sensory loss through tactile sensations evoked by directly activating cortex electrically^5–9^ (**Figure 1A-C**). When combined with a motor iBCI, ICMS-feedback can significantly improve hand and arm function^5^. Beyond somatosensation, ICMS may also restore other lost senses, such as vision for patients with retinal or subcortical visual impairments^10,11^. Unfortunately, no clinically available ICMS devices exist, in part due to an absence of evidence of safety and longevity for ICMS in humans, with the previous longest study lasting only 4 months^22^. While several studies have addressed microelectrode array recording stability^12–15^ and documented ICMS longevity in humans up to three years^14^, safety studies are extremely limited. Studies in rodents^16^, cats^17–19^, and macaques^20,21^ have been performed, but rarely extended beyond a year. For more than a decade, we have conducted a multi-site early feasibility trial under an FDA investigational device exemption. One goal of this study was to evaluate the safety of a bidirectional iBCI for neural recording and stimulation with the ultimate objective of restoring sensorimotor function of the arm and hand to people with SCI. As part of this clinical trial, we have used ICMS for extensive psychophysical^5,23–26^ and functional^27–30^ testing across five participants. Here we assess the long-term viability of ICMS by performing a retrospective analysis on the safety, efficacy, and longevity of ICMS spanning 24 cumulative years and over 168 million ICMS pulses (**Figure 1D**).

**Figure 1.**
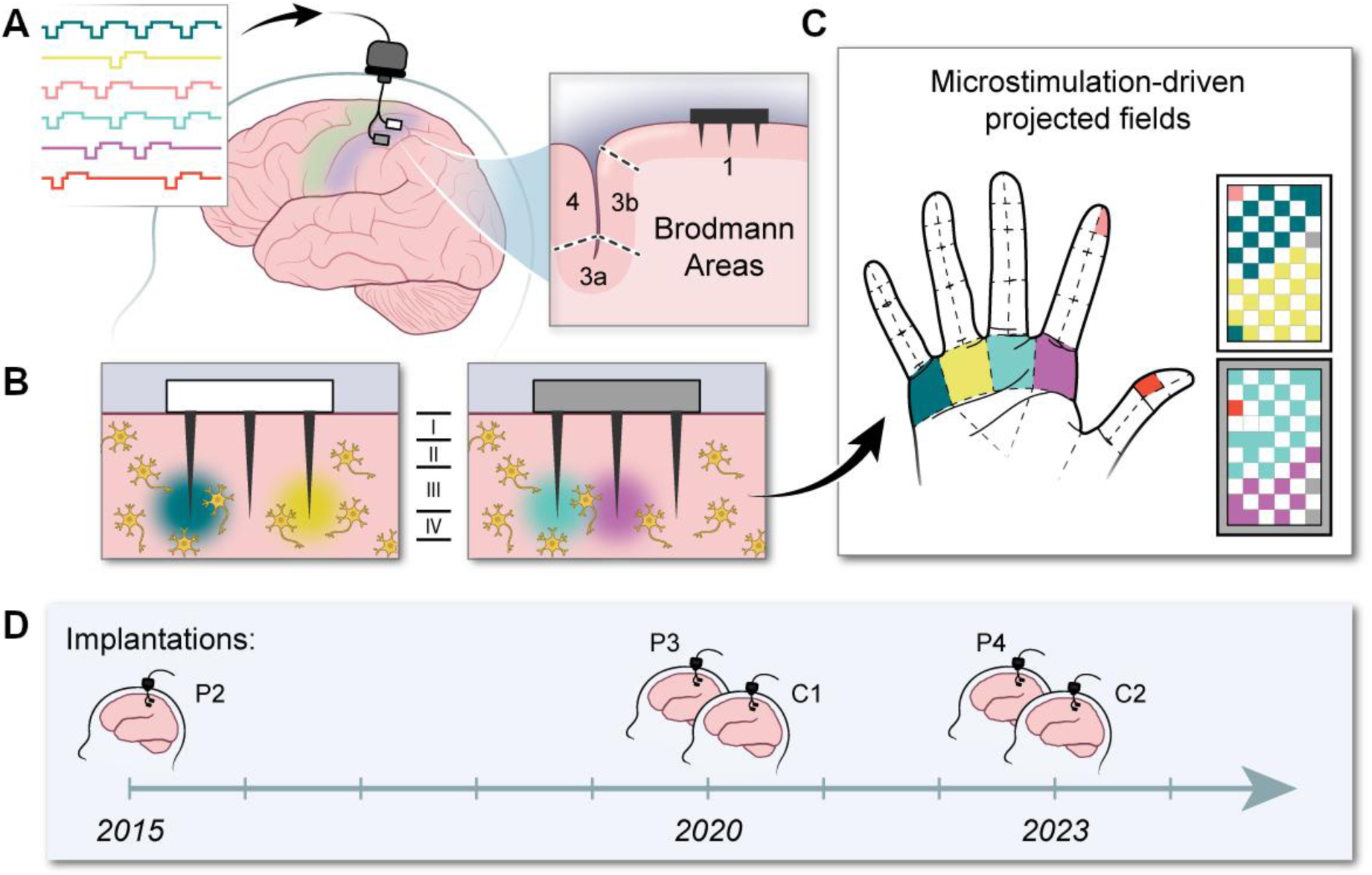
Clinical trial setup and overview. **A|** Two Blackrock NeuroPort intracortical microelectrode arrays were placed in Area 1 of somatosensory cortex (shaded blue) targeting layer IV. ICMS pulses (illustrated in left inset) were delivered to the arrays via a CereStim R96 stimulator. A cross-section of the NeuroPort arrays implanted into Area 1 is shown in the right inset. **B|** ICMS delivered through the arrays activates neurons near the electrode tip. **C|** This activation evokes sensory percepts on the hand in a somatotopically organized manner. Colors on the microelectrode arrays denote regions on the hand where ICMS evokes conscious percepts. **D|** Timeline of implants for all participants in the clinical trial.

## Methods

### Trial design and oversight

This clinical trial includes participants at both the University of Pittsburgh and the University of Chicago. The study is conducted under an investigational device exemption from the US Food and Drug Administration, institutional review board protocols at both sites, and details of the trial are listed on clinicaltrials.gov (NCT01894802). Safety data and scientific progress were presented bi-annually to an independent data and safety monitoring board composed of experts in the field. All participants described here remain implanted at the time of writing (Supplementary Appendix).

### Trial procedures

Participants were each implanted with two Blackrock NeuroPort microelectrode arrays (Blackrock Neurotech Inc., Salt Lake City, UT, USA) in the hand representation of Brodmann’s Area 1 and another two arrays in the hand and arm region of motor cortex contralateral to their dominant hand (all right-handed)^31^. ICMS was delivered with a CereStim R96 (Blackrock Neurotech Inc., Salt Lake City, UT, USA). Stimulation levels depended on experimental needs and were not intentionally varied for the purposes of this paper.

### Safety assessments

The primary outcome measure for the clinical trial is the safety of the participants, specifically, that the device does not need to be explanted due to a safety concern within the 1^st^ year of implantation. This <u>criterion was met for all participants</u>. Here, we describe device-related adverse events (AEs) that occurred directly as a result of ICMS. These consisted entirely of sensations that persisted beyond the duration of ICMS – known as persistent sensations – which participants were instructed to report throughout the clinical trial. While no adverse clinical sequalae were observed due to these persistent sensations, they were logged as AEs along with stimulus parameters and any other sensory effects.

### Efficacy assessments

Electrodes were determined to be functional if their median detection threshold over a 6-month period was below 100 μA. The proportion of each participant’s hand that was covered by the projected fields (the location where ICMS evoked a sensation on the hand) at each timepoint was computed as the union of the functional electrodes projected fields^28^. Finally, the stability of the sensory qualities evoked by each electrode was computed by measuring the average pairwise correlation coefficient between quality reports over time.

### Longevity assessments

As efficacy assessments were collected routinely throughout the study, longitudinal assessments were made by measuring changes in detection threshold for each electrode and in the total projected field coverage. Additionally, we examined changes in electrode health metrics including signal-to-noise ratio, peak-to-peak voltage, interphase voltage, and impedance.

### Data and code availability

Code for data processing and analysis can be found on GitHub (https://github.com/CorticalBionics/ICMS_Safety). Deidentified data generated in this study have been deposited in the Data Archive BRAIN Initiative repository (https://dabi.loni.usc.edu/dsi/LVUPBGAXILJ6).

## Results

### Trial population

Five participants were implanted between 2015 and 2023. All were male, >25 years of age at the time of implantation, right-handed, and had varying degrees of residual sensation on the hand contralateral to the implant. Participants were implanted for 26-122 months and completed 191-1357 experimental sessions, 42-78% of which included ICMS that culminated in a total of 588 hours of stimulation, 168 million pulses, and 1666 millicoulombs of charge (**Table 1** and **Supplementary Figure 1**).

**Table 1.** Stimulation characteristics for each participant.

| Characteristic | Participant |  |  |  |  |
| --- | --- | --- | --- | --- | --- |
|  | C1 | C2 | P2 | P3 | P4 |
| **Months implanted** | 56 | 26 | 122 | 59 | 27 |
| **Total sessions** | 650 | 300 | 1357 | 295 | 191 |
| **Stimulation sessions** | 504 | 149 | 1023 | 189 | 80 |
| Per month | 9.0 | 5.7 | 8.4 | 3.2 | 3.0 |
| **Duration** |  |  |  |  |  |
| Total (hours) | 308.8 | 44.8 | 156.4 | 65.7 | 12.1 |
| Per session (minutes)* | 23.4<br>(13.0 - 40.7) | 14.4<br>(6.7 - 25.1) | 4.1<br>(1.3 - 10.9) | 12.8<br>(6.4 - 24.6) | 6.1<br>(3.5 - 13.3) |
| **Trials (thousands)** |  |  |  |  |  |
| Total | 227.9 | 49.3 | 211.3 | 68.6 | 19 |
| Single-electrode (%) | 170.6 (75%) | 30.5 (62%) | 183.9 (87%) | 50.2 (73%) | 12.5 (66%) |
| Multi-electrode (%) | 57.3 (25%) | 18.8 (38%) | 27.4 (13%) | 18.4 (27%) | 6.5 (34%) |
| **Pulses** |  |  |  |  |  |
| Total (millions) | 66 | 35.9 | 51.7 | 11.8 | 2.7 |
| Per session (thousands)* | 97.3<br>(51.9 - 192.6) | 96.1<br>(38.5 - 253.6) | 38.7<br>(15.8 - 64.1) | 51.3<br>(28.8 - 86.9) | 26.7<br>(17.4 - 41.4) |
| Per electrode (thousands)* | 682.6<br>(404.7 - 1216.4) | 528.1<br>(332.2 - 793.9) | 400.4<br>(214.5 - 810.4) | 119.1<br>(69.0 - 233.1) | 32.4<br>(25.6 - 51.8) |
| **Charge** |  |  |  |  |  |
| Total (mC) | 581.3 | 508.2 | 462.1 | 93.5 | 20.8 |
| Per session (mC)* | 0.9 (0.4 - 1.6) | 1.2 (0.5 - 3.0) | 0.3 (0.1 - 0.6) | 0.4 (0.2 - 0.8) | 0.2 (0.1 - 0.3) |
| Per electrode (mC)* | 6.46<br>(3.8 - 12.4) | 7.84<br>(4.4 - 10.7) | 3.55<br>(1.6 - 7.1) | 0.91<br>(0.4 - 2.0) | 0.22<br>(0.2 - 0.4) |
| Per phase (nC)* | 9.12<br>(8.5 - 10.0) | 14.25<br>(13.3 - 15.0) | 8.41<br>(7.1 - 9.1) | 7.56<br>(6.5 - 8.3) | 7.10<br>(6.4 - 7.7) |

\* Median and interquartile range.

### Safety

There were 53 ICMS-related AEs (3–25 per participant, **Table 2**), all of which were categorized as persistent sensations: those that persist beyond the end of the stimulus train. Sensations always remained localized to the region of the original ICMS-evoked percept, no medical intervention was ever required, and there were no lasting effects. Despite the minimal clinical repercussions of persistent sensations, they are still considered AEs due to their unintended nature and the potential complications they could cause when delivering feedback (see Supplementary Appendix). Persistent sensations were extremely infrequent across all participants: on average, one persistent sensation for every 22,000 stimulation trials. These sensations predominantly lasted for less than 10 seconds, while the maximum reported persistent sensation lasted for approximately 9.5 minutes. The persistent sensations were typically described as continuations of the ICMS-evoked sensation (i.e. tingling or buzzing) and were never described as painful. Persistent sensations were more common following multi-electrode stimulation for 3 of the 5 participants, implying that either the activated volume of cortex or instantaneous injected charge may partially determine their likelihood. The consensus opinion is that persistent sensations reflect the perceptual consequence of stimulus-induced after-discharges^32^ (see Discussion). Non ICMS-related AEs are discussed in the Supplementary Appendix.

**Table 2.** Summary of persistent sensations for each participant.

|  | Participant |  |  |  |  |  |  |  |  |  |
| --- | --- | --- | --- | --- | --- | --- | --- | --- | --- | --- |
|  | C1 |  | C2 |  | P2 |  | P3 |  | P4 |  |
| Total | 3 |  | 3 |  | 25 |  | 16 |  | 6 |  |
| Duration |  |  |  |  |  |  |  |  |  |  |
| 0-10 s | 3 | 100% | 0 | 0% | 21 | 84% | 3 | 19% | 6 | 100% |
| 10-120 s | 0 | 0% | 3 | 100% | 4 | 16% | 12 | 75% | 0 | 0% |
| 120 s-10 min | 0 | 0% | 0 | 0% | 0 | 0% | 1 | 6% | 0 | 0% |
| Single/Multi Electrode |  |  |  |  |  |  |  |  |  |  |
| Single | 1 | 33% | 0 | 0% | 10 | 36% | 12 | 75% | 3 | 50% |
| Multi | 2 | 67% | 3 | 50% | 15 | 55% | 4 | 25% | 3 | 50% |
| Frequency |  |  |  |  |  |  |  |  |  |  |
| Sessions/PS | 168.0 |  | 49.7 |  | 40.9 |  | 11.8 |  | 13.3 |  |
| kTrials/PS | 76 |  | 16 |  | 8 |  | 4 |  | 3 |  |
| k SE Trials/PS* | 170.6 |  | - |  | 18.4 |  | 4.2 |  | 4.2 |  |
| k ME Trials/PS† | 28.7 |  | 6.3 |  | 1.8 |  | 4.6 |  | 2.2 |  |

\* Single-electrode kilo-trials. † Multi-electrode kilo-trials. PS= persistent sensation

Four of the five participants retained at least partial tactile sensation in regions which the electrodes evoked sensations. This provided another measure of ICMS safety, as stimulation-induced tissue damage could increase these natural sensory thresholds^28^. We used monofilaments to measure tactile thresholds before implantation and at the time of writing and found no consistent change (**Supplementary Table 1**), suggesting that neither the implantation nor ICMS had an effect on natural sensation.

### Efficacy and longevity

Our primary metric for ICMS efficacy was single-electrode detection thresholds, as this confirms that electrical current is successfully being delivered to the tissue, the electrode is intact, and there is excitable neural tissue near the electrode tip. Across participants, we completed 118-2032 measurements, which were regularly assessed since the time of first sensation (14-111 days after implantation, **Figure 2A, Supplementary Figure 2A**). Initial thresholds were low for four of the five participants (median range: 14.5-22.5 μA), while the fifth participant exhibited higher thresholds (median: 61.5 μA, and also had the longest time to first sensation). We computed the overall change in the detection threshold as a function of time for each electrode and observed an average 3.54±11.52 μA increase per year, though this varied substantially across participants (**Figure 2B**, one-way ANOVA: Participant[4], F=3.33, p=0.0113). Since detection thresholds can only be measured if they are below the stimulation limit, these values only reflect changes for functioning electrodes (i.e. those that can be detected). Therefore, we also quantified the number of functioning electrodes over time and found that >75% of electrodes for three of the five participants were functional one year after implantation (**Figure 2C**). This proportion decreased over time: after 4 years, 79% and 75% of Participants C1 and P3 electrodes were functional, while after 10 years, 54% of Participant P2’s electrodes were functional. Participant P4 demonstrated accelerated increases in detection threshold while Participant C2’s thresholds (which were initially higher) showed improved sensitivity. At the most recent timepoint, 43% and 57% of P4’s and C2s electrodes were functional, respectively. As we preferentially stimulated low threshold electrodes (**Supplementary Figure 2B**, Spearman correlation: p < 0.05 for 3/5 participants), we assessed if the total charge delivered during the study influenced the change in threshold and found no relationship (**Supplementary Figure 2C**).

**Figure 2.**
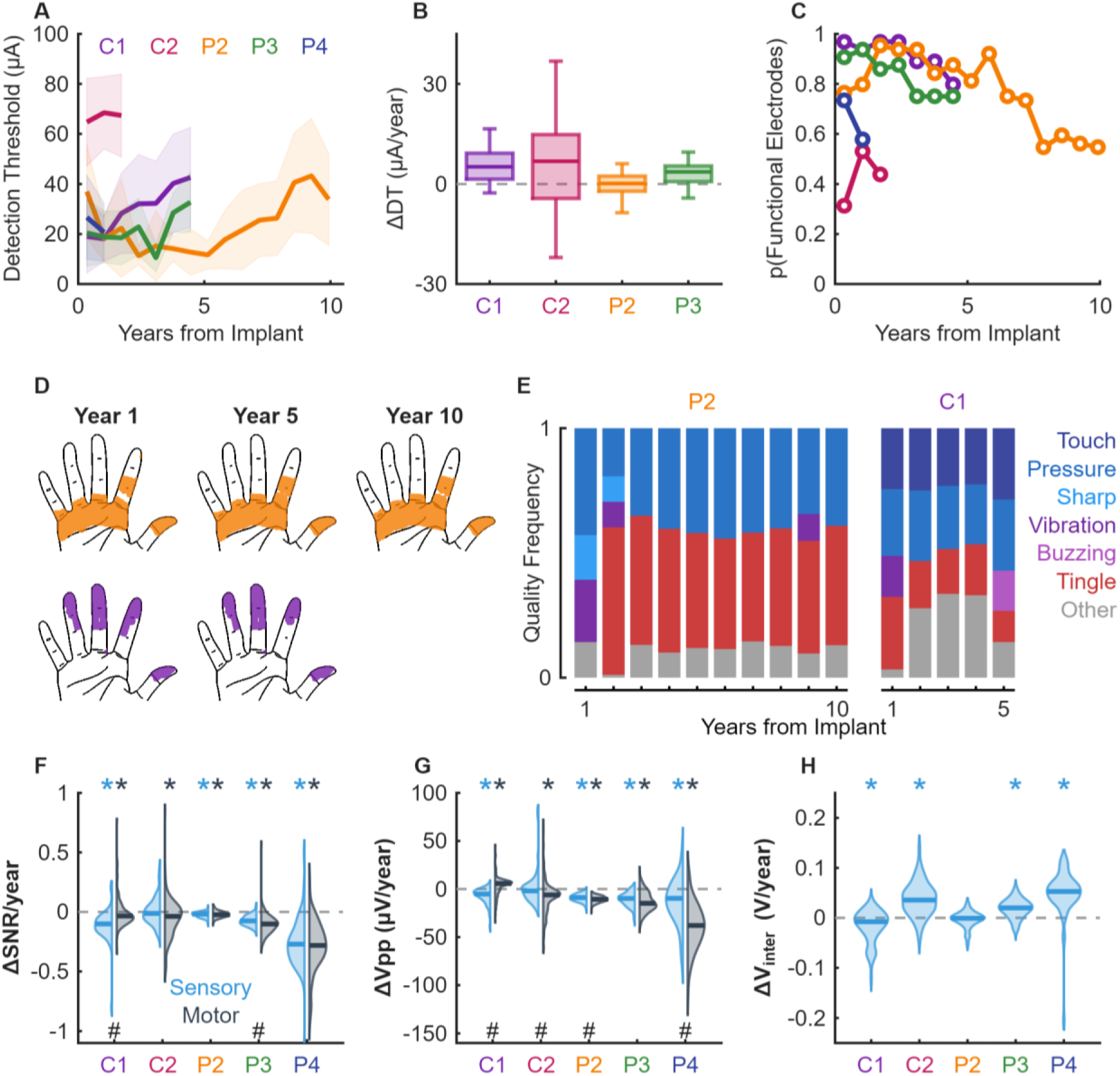
Changes in perceptual and signal quality metrics over time. **A**| Detection thresholds over time across electrodes for each participant. Mean and standard deviation indicated with a line and shaded area respectively. **B|** Distribution of linear regression slopes fitted to the detection thresholds over time for each electrode and participant. Box and whiskers indicate 5^th^, 25^th^, 50^th^, 75^th^, and 95^th^ percentiles. Participant P4 not shown due to insufficient data. **C|** Proportion of functional electrodes for each participant determined by whether or not their detection threshold is above or below 100 μA. **D|** Example hand coverage plot for Participants C1 and P2 over time. **E|** Distribution of sensation qualities reported by the same participants in **D** over time. The category ‘Other’ represents the summed frequency of qualities that are individually represented in less than 10% of the surveys. **F|** Distribution of linear regression slopes computed for the SNR of each electrode over time, split by motor and sensory arrays. Asterisks above indicate that the distribution of the array significantly deviated from 0 using a sign test with Holm-Bonferroni correction. Hashes below indicate a significant difference (p < 0.05) between the motor and sensory array slopes for the participant using a Wilcoxon rank-sum test with Holm-Bonferroni correction. **G|** Same as **F** but for peak to peak voltage. **H|** Distribution of linear regression slopes computed for the interphase voltage of each electrode over time for the sensory arrays.

Examining perceptual changes in ICMS-evoked sensations over time, we sought to quantify somatotopic coverage by analyzing survey responses (**Supplementary Figure 3A**). Computing the relative coverage over time (**Figure 2D, Supplementary Figure 3B**), we found that despite the reduction in the proportion of functional electrodes, the somatotopic coverage remained largely unchanged due to spatially-redundant projected fields. We next examined the stability of the qualitative reports over time (**Figure 2E**). Sensations tended to be described as mechanical (e.g., pressure, touch) or paresthetic (e.g., tingle, electrical), with a minority representing other sensations (e.g., vibration, buzzing)^33^. Quality reports were reliable for all electrodes for Participants C1, P2, and P3 (**Supplementary Figure 3C**, Pearson’s correlation with Holm-Bonferroni correction: *r*>0.81, p<0.006), but not for Participants C2 and P4 (r=0.65 and -0.13, p>0.05), both of whom had higher detection thresholds. Additionally, we found that the reported naturalness of the sensations remained stable or increased over time. However, the naturalness rating began decreasing for Participant P2 after the 8^th^ year (**Supplementary Figure 3D**). Finally, examining ICMS-associated pain or discomfort, we found that while two participants regularly reported mild pain (**Supplementary Figure 3E**), the pain was substantially less than any baseline level associated with their injury (median: 1.5 vs 7, **Supplementary Figure 3F**, Wilcoxon rank-sum test: p<0.05 for both) indicating that the ICMS-related pain reports were likely biased by their general discomfort.

Regarding other potentially long-lasting effects of ICMS, we investigated neural recording quality by measuring the signal-to-noise ratio (SNR), peak-to-peak voltage (V_pp_), and 1 kHz impedance, reasoning that decreases in any metric that were greater on the stimulated electrodes could reflect electrode-tissue interface degradation. We found that both SNR and V_pp_ tended to decrease for both the sensory and motor electrodes, except for Participant C1, in which V_pp_ *increased* slightly over time for the motor electrodes (SNR: -0.28 to -0.02/year, V_pp_: -32.27 to 0.8 μV/year, **Figure 2F,G**). Comparing motor and sensory electrodes, only two of the five participants had significant differences in ΔSNR, and in opposite directions (Wilcoxon rank-sum test: p<0.05, **Figure 2F, Supplementary Figure 4A**). For ΔV_pp_, four participants had statistically different changes between the motor and sensory arrays with the sensory electrodes experiencing a slower decline in V_pp_ than the motor electrodes in three participants (**Figure 2G, Supplementary Figure 4B**). Impedances decayed exponentially for all electrodes as previously observed^12^ (**Supplementary Figure 4C**) with no consistent differences between stimulated and unstimulated electrodes. Additionally, we assessed the interphase voltages over time; three of the five participants demonstrated significant, though inconsistent changes (**Figure 2H**, **Supplementary Figure 4D**, -0.025 to 0.070 V/year, Sign-test, p<0.05).

Finally, we examined whether the quantity of stimulation affected signal quality. Using the correlation between the charge delivered to each electrode and the respective change in SNR, V_pp_, or interphase voltage, we found no consistent relationships across participants, implying that changes in these metrics were due to time and not the quantity of stimulation (**Supplementary Figure 5A**). Examining electrode detectability specifically, we found that neither absolute signal quality values nor their relative changes were indicative of changes in ICMS detectability (**Supplementary Figure 5B-H**) and that the spatial distribution of detectable electrodes was consistent over the duration of the implants (**Supplementary Figure 6**).

## Discussion

The data presented here represents the largest safety study of ICMS in humans to date. ICMS was both safe and effective for up to 10 years. While detection thresholds increased over timescales that were similar to reductions in electrode health metrics, there were no robust relationships between these observations across participants. Moreover, we found no relationship between the amount of stimulation delivered to each electrode and any measure of performance or longevity, although the differences in total charge and implant duration across different participants may affect our ability to detect changes. Ultimately, while performance slowly decreased with time, the changes appear more related to implant duration than ICMS delivery. The main AE that was observed using ICMS was the infrequent occurrence of brief persistent sensations across the participants, a modest risk relative to that associated with surgery and implantation (see Supplementary Appendix).

Animal studies using microelectrodes, particularly in cats, demonstrated that ICMS could lead to cell death in the immediate vicinity of the electrode tips^17–19^. More recent chronic studies with refined stimulation parameters focused on limiting duty cycle found no effect of stimulation^20^ and defined the safety limits for human studies. Since then, minimal long-term safety data on ICMS in humans have been presented, even though several studies have broadly established the ability of ICMS to evoke artificial tactile percepts in at least 10 people^7,8,14,31^, with this current clinical trial representing the longest and most populous study to date.

While ICMS was consistently safe, cross-participant comparisons highlight substantial variation in terms of efficacy. Participants P2 and C2 did not report sensations until four and 16 weeks after implantation respectively, while all other participants reported sensations on their first stimulation session (∼2 weeks). The reason for this may be related to residual, post-surgical tactile sensations ‘masking’ ICMS-evoked sensations^5^. The most variability occurred with detection thresholds. For Participant C2, initially high detection thresholds decreased somewhat, but still remain higher than other participants. Participant P2, who has been implanted for 10 years, had low detection thresholds for approximately 6 years before thresholds began increasing. These differences may arise from differences in electrode depth, residual sensation, or other factors, though the exact reasons are unknown.

ICMS is comparatively safe among neuromodulation approaches. In DBS trials, stimulus-induced AE rates represent approximately 23% of the total AEs^34^ and one study of surface stimulation for visual restoration resulted in approximately one stimulus-induced AE per four stimulation sessions^35^. Critically, the nature of the ICMS-evoked AEs is significantly less severe than those described for other neuromodulation approaches. Nevertheless, it must be recognized that we delivered substantially less charge than a clinical device typically would, amounting to only ∼20 minutes of stimulation per session, several times a week. While real world usage will vary, we would expect >10x more stimulation to be appropriate^36^. Similarly, the primate safety study which preceded this clinical trial^20^ delivered over 10,000 millicoulombs of charge over 6-months to each animal, considerably more than we delivered to all participants combined. The data presented here imply that more stimulation is unlikely to be detrimental to safety or longevity, and studies on explanted electrodes^14^ support this.

An important caveat is that we lack an understanding of the nature and etiology of ICMS-evoked persistent sensations. Prolonged or intensifying sensations may raise concern for paroxysmal, synchronous, and self-limited electrophysiological activity known as afterdischarges^32^. Importantly, afterdischarges are not clinical seizures; they are brief, localized discharges commonly observed following electrical stimulation and do not typically propagate or include behavioral manifestations. Indeed, no lasting effects were observed in any participants following the persistent sensation events.

A challenge in further studying persistent sensations is the extremely low occurrence rate. Critically, within the 100 μA and 300 Hz stimulation limits we explored; no combination was found that reliably induced persistent sensations. However, we observed that multi-electrode stimulation may be more likely to evoke them. Given that recent work has shown that multi-electrode stimulation significantly improves performance in functional tasks^28^ and will likely be needed to convey complex object information^29^, we will need to balance safety implications with functional benefits. Conversely, multi-electrode stimulation reduces the stimulus current on individual electrodes required to detect a stimulus^28,37^, perhaps enabling effective stimulation at lower per-electrode intensities.

Importantly, all our participants are screened for evidence of seizure disorders prior to device implantation, stimulus parameter limits have been defined by large datasets in animals^20^, and stimulus sessions are monitored closely with clearly defined protocols for intervention. Improved hardware that facilitates low-noise simultaneous recording during ICMS will allow evaluation of neural activity associated with persistent sensations, providing deeper neurophysiological insight. This may enable real-time early detection systems to alter or stop stimulation entirely before a persistent sensation is evoked.

In conclusion, we find no serious AEs associated with long-term ICMS in the somatosensory cortex; the few persistent sensations that did occur resolved quickly and without clinical concern. While this study was limited to stimulation of somatosensory cortex, we believe that the safety profile is likely to extend to other regions, though definitive clinical trials are still necessary. Regarding longevity, while functionality decreased, the fact that ICMS-evoked sensations were stable over up to 10 years in terms of both quality and location^28^ indicates that ICMS can be used as a valuable tool for extended durations.

## Data Availability

All code for data processing and analysis can be found on GitHub (https://github.com/CorticalBionics/ICMS_Safety). The deidentified data generated in this study have been deposited in the Data Archive BRAIN Initiative (DABI) under the project code LVUPBGAXILJ6 (https://dabi.loni.usc.edu/dsi/LVUPBGAXILJ6).

https://dabi.loni.usc.edu/dsi/LVUPBGAXILJ6

## Supplementary Appendix

### Participants

All participants provided written consent before any study procedures were conducted and were reconsented following protocol modifications. Participants were recruited using a variety of strategies including research registries, advertisements, and clinician referrals. All participants had limited or no ability to use one or both hands due to a cervical SCI that occurred a minimum of 1 year before enrollment in the study. Due to the nature of the clinical trial, randomization was not possible.

Participant C1 (male), 55-60 years old at the time of implant, presented with a C4-level ASIA D SCI that occurred 35 years prior. Monofilament sensory tests revealed spared deep sensation but diminished light touch in the right hand (detection thresholds ranged from 0.6 to 2.0 g across digit tips).

Participant C2 (male), 60-65 years old at time of implant, presented with a C4-level ASIA D SCI, along with a right brachial plexopathy, that occurred 4 years prior. He was nearly insensate across the whole right hand (monofilament sensory thresholds were 15-60 g across digits 2-5 and >300 g on D1).

Participant P2 (male), 25-30 years old at the time of implant, presented with a C5 motor/C6 sensory ASIA B SCI that occurred 10 years prior to implant. He was insensate in the ulnar region of the hand (digits 3-5) on both the palmar and volar surfaces but retained both diminished light touch and deep sensation on the radial side (digits 1-2) (monofilament sensory thresholds were 1.4 g to 8 g on the thumb and index, respectively, and 180 g on the middle finger).

Participant P3 (male), 25-30 years old at the time of implant, presented with a C6 ASIA B SCI that occurred 12 years prior. He was insensate in the ulnar region of the hand on both the palmar and volar surfaces but retained diminished light touch and deep sensation on the radial side (monofilament sensory thresholds were 0.07 g and 1.6 g on the thumb and index and 8 g on the middle finger).

Participant P4 (male), 30-35 years old at the time of implant, presented with a C4 ASIA A SCI that occurred 11 years prior. He had trace muscle activity in his right bicep but is otherwise paralyzed below the neck with no spared sensation at or below the C5 dermatome.

### Implantation and related adverse events

The surgical implantation of intracortical microelectrode arrays was well tolerated across all five participants, with no major intraoperative or postoperative complications observed. There were no instances of infection, intracranial hemorrhage, neurological deterioration, or new-onset seizures attributable to the implantation procedure. Two participants required a minor surgical revision due to a loose percutaneous pedestal, which was successfully resecured without complication. In addition, minimal skin debridement was performed in one of those participants at the pedestal site to maintain skin integrity. These isolated events were not associated with any neurological sequelae, systemic infection, or electrode malfunction. Overall, the implantation procedures were deemed safe, and participants recovered without significant adverse outcomes directly attributable to the surgical placement of the devices.

### Microelectrode arrays and ICMS

Each NeuroPort electrode array for ICMS consisted of a 6 x 10 grid of electrodes with a 400 μm interelectrode spacing. 32 of the 1.5 mm long electrodes were available for stimulation and all electrodes were coated with sputtered iridium oxide to increase charge injection capacity and improve stimulation ability^38^. Stimulation pulses were current-controlled, biphasic, and charge-balanced, consisting of a 200 µs cathodic leading phase, a 100 µs interphase period, and a 400 µs half-amplitude anodic phase. The voltage transient for each stimulus pulse was recorded and logged in real time through a previously described custom recording system^5^.

### Persistent sensations

Despite the lack of side effects, we consider persistent sensations as AEs because of the potential for a prolonged, expanding, or strengthening persistent sensation to be indicative of seizure activity. Moreover, persistent sensations are undesirable from a functional perspective as they will confer a false sense of contact or force to the participants when they occur. Critically, many people with SCI report ongoing or intrinsic spontaneous sensations as a result of their injury^39^ – similar to phantom limbs – and as such participants would indicate on some occasions that they could not differentiate between these sensations and ICMS-induced persistent sensations. In the interest of safety, participants were encouraged to report a persistent sensation on these occasions. Consequently, the persistent sensations reported here may include some false positives.

If a persistent sensation lasted less than 10 seconds, no additional action was taken. If a persistent sensation lasted 10 s to 2 min, stimulation was stopped on that electrode for the day. If a persistent sensation lasted between 2 min and 10 min, stimulation on all electrodes was stopped for the day. A study physician was notified immediately for any persistent sensation lasting more than 2 min.

### Manufacturer involvement

Blackrock Neurotech Inc., the manufacturer of the device, was consulted extensively for the development of the original Investigational Device Exemption from the FDA, including providing access to implant and external component safety, biocompatibility, and other required data. Further, they were consulted regarding implantation procedures for some participants. Blackrock Neurotech has assisted with applications for federal grants and has provided limited funding for specific aspects of this trial. They have been given access to device safety data but were not consulted for this publication and have never placed any scientific restrictions on publication of data generated with the device.

### Detection thresholds

For psychophysical estimation of the single-electrode current amplitude detection thresholds, we used the method of constants or the 3-down 1-up transformed staircase, both of which target ∼50% detection threshold. Pulse frequency and train duration were held constant (100 Hz, 1 second) while the current amplitude varied. All trials used a 2-interval forced choice paradigm such that each trial contained a ‘catch’ interval with no stimulus and a ‘test’ interval with a stimulus. The interval containing the ‘test’ stimulus was randomly selected on each trial and the participant was asked to report the interval that they believed contained the stimulus.

### Projected field mapping

To assess the locations of the projected fields evoked by each electrode, we used the pixel coordinates reported on a 2D digital representation of the hand following repeated 100 Hz, 60 μA, 1 second stimulus trains. We combined all survey reports over the implant duration and report the pixels that were selected by the participants on at least 30% of surveys as previously reported^28^. The quality of the evoked percept was assessed using a survey that measured the perceived intensity (scale of 0-10, ranging from low to high) of various mechanical, movement, and paresthetic descriptors^33^. In addition, participants were asked to rate the experienced pain and naturalness of each sensation. This survey was repeated every one to three months for each enabled stimulus electrode.

### Impedance measurements

Electrode impedance measurements were taken at 1 kHz using Central (Blackrock Neurotech, UT, USA) at both the beginning and end of each session. Values within session were averaged. Note that one motor array in Participant C1 and one sensory array in Participant C2 had uncharacteristically low impedances, and were removed from further analysis. Both arrays had median electrodes impedances of 32 kOhm.

### Interphase voltage monitoring

To assess interphase voltage stability, we delivered a 10 μA, 100 Hz, 0.5 second long stimulus to each electrode at the beginning of each stimulation session^5^. Stimuli were delivered to up to 12 electrodes simultaneously, distributed spatially across both arrays to minimize potential voltage field interactions. The voltages associated with these current pulses were sampled at 100 kHz, and we extracted the last voltage value prior to the onset of the anodic phase of the waveform as a metric of the interphase voltage to assess changes in electrode integrity^40^. Because the voltages are recorded prior to an inline safety capacitor in the stimulator, we report changes in interphase voltage as the absolute values represent the combined voltage across the capacitor and microelectrode.

### Statistical analyses

All analyses were retrospective and thus no power calculations were performed prior to analyses. Holm-Bonferroni post-hoc correction was used whenever multiple comparisons were made unless explicitly stated otherwise.

Stimulus duration was computed by subtracting the time between the first and last pulse on each trial. In cases where there were gaps in stimulation within a trial greater than 200 ms, for example an inter-stimulus interval or stimulation below 5 Hz, these periods were excluded. To compute the current delivered on each session, we summed the commanded currents for each pulse and electrode.

To compute the changes in electrode detection threshold over time for each electrode, we used linear regression between the number of days since implant of each threshold test and the measured threshold on each day. Regressions were only performed if at least 5 threshold measurements were made for each electrode for robustness. To then compare slopes between participants, a 1-way ANOVA was used on the computed slopes. To determine the proportion of functional electrodes, we grouped threshold measurements for each electrode by the time after implantation (bin width = 250 days). Then, for each electrode we took the median threshold for all measurements in each time bin. If no measurements were taken for a single bin for whatever reason, the mean value of the adjacent bins was used. Electrodes were considered ‘non-functional’ if either no threshold measurements had been made during the 250-day period or over 50% of detection threshold measurements were unsuccessful.

To compute projected field coverage over time, we computed the projected fields of each electrode as any pixel that was reported on at least 33% of surveys during the first year^28^. The total coverage was then represented by the union of all pixels across electrodes. To determine the relative coverage over time, we recalculated the union using the appropriate functional electrodes at each time point before normalizing the area by the timepoint at which maximum coverage was observed.

To measure the qualities associated with each electrode, 1-second-long pulses at 60 μA and 100 Hz were delivered to each electrode. Participants were given a list of verbal descriptors that they could select from and give a value between 0 and 10 for each descriptor. Participants were allowed to request unlimited repetitions of each stimulus to maximize their reporting certainty. In this same survey, participants were asked to rate both the overall naturalness of the sensation and any pain associated with it each on a scale between 0 and 10. Electrodes that evoked a sensation less than 30% of the time, as well as qualities that were never reported in response to stimulation, were excluded from analysis. To determine if the quality reports were consistent for each electrode, we computed the Pearson correlation between all pairs of surveys for a given electrode and computed the average correlation for pairs within 1-year bins. These correlations were then compared to shuffled controls where within each electrode the quality values were shuffled before the correlation was measured.

Electrode health measurements included the signal-to-noise ratio (SNR) and peak-to-peak voltage (V_pp_). These metrics were extracted from daily ‘baseline’ datasets that consisted of one minute long recordings that were performed at the beginning of every session for each participant. Before recordings began, a 4.5x root mean square threshold was applied to each electrode and waveforms were captured at 30 kHz whenever the threshold was crossed. To compute the SNR, the noise was computed as 3 times the standard deviation of the first 5 samples of each waveform (166 μs). The waveforms were then averaged, and the signal was computed as the maximum absolute amplitude and the SNR was then computed as the signal divided by the noise for each electrode. The V_pp_ was assessed as the range between the maximum and minimum values of the averaged waveform. To compare metrics between sensory (stimulated) and motor (unstimulated) arrays, linear regression was used to compute the relationship between time and the relevant metric (slope). The slopes were then split depending on whether they belonged to motor or sensory arrays. To assess if the slopes were consistently positive or negative, a sign-test was used. To compare the slopes between arrays, a Wilcoxon rank sum test was used.

## Acknowledgements

We would like to thank the study participants for their generous contribution to the advancement of science. The work at the University of Chicago and University of Pittsburgh was supported by the National Institute of Neurological Disorders and Stroke of the National Institutes of Health under Award Numbers UH3 NS107714, R35 NS122333, U01 NS108922, U01 NS123125, and R01 NS130302 and by the Defense Advanced Research Projects Agency and Space and Naval Warfare Systems Center Pacific under Contracts N66001-16-C-4051 and N66001-10-C-4056. TGH and AHA are supported by the NDSEG National Defense Science and Engineering Graduate Fellowship. JY is supported by Department of Energy Computational Science Graduate Fellowship. RL is supported by the Walter-Benjamin Program fellowship from the German Research Foundation. We would like to thank Sliman Bensmaia for his instrumental involvement in the clinical trial as well as Carleigh May for her support in clinical coordination of the study as well as all past and present members of the team who collected any amount of data used in the retrospective analyses presented here.

## Disclosures

RG is on the scientific advisory board of NeuroWired and has consulted for Blackrock Neurotech. RG, JC, and MB have received research funding from Blackrock Neurotech. CMG has received sponsored travel from Blackrock Neurotech. JMW consults for Blackrock Neurotech. GV holds shares of “MYNERVA AG” and serves as a consultant for NeuroOne Medical Technologies Corporation. JGM is a consultant for DIXI Medical. The remaining authors declare no competing interests.

## Supplementary Figures & Tables

**Supplementary Figure 1.**
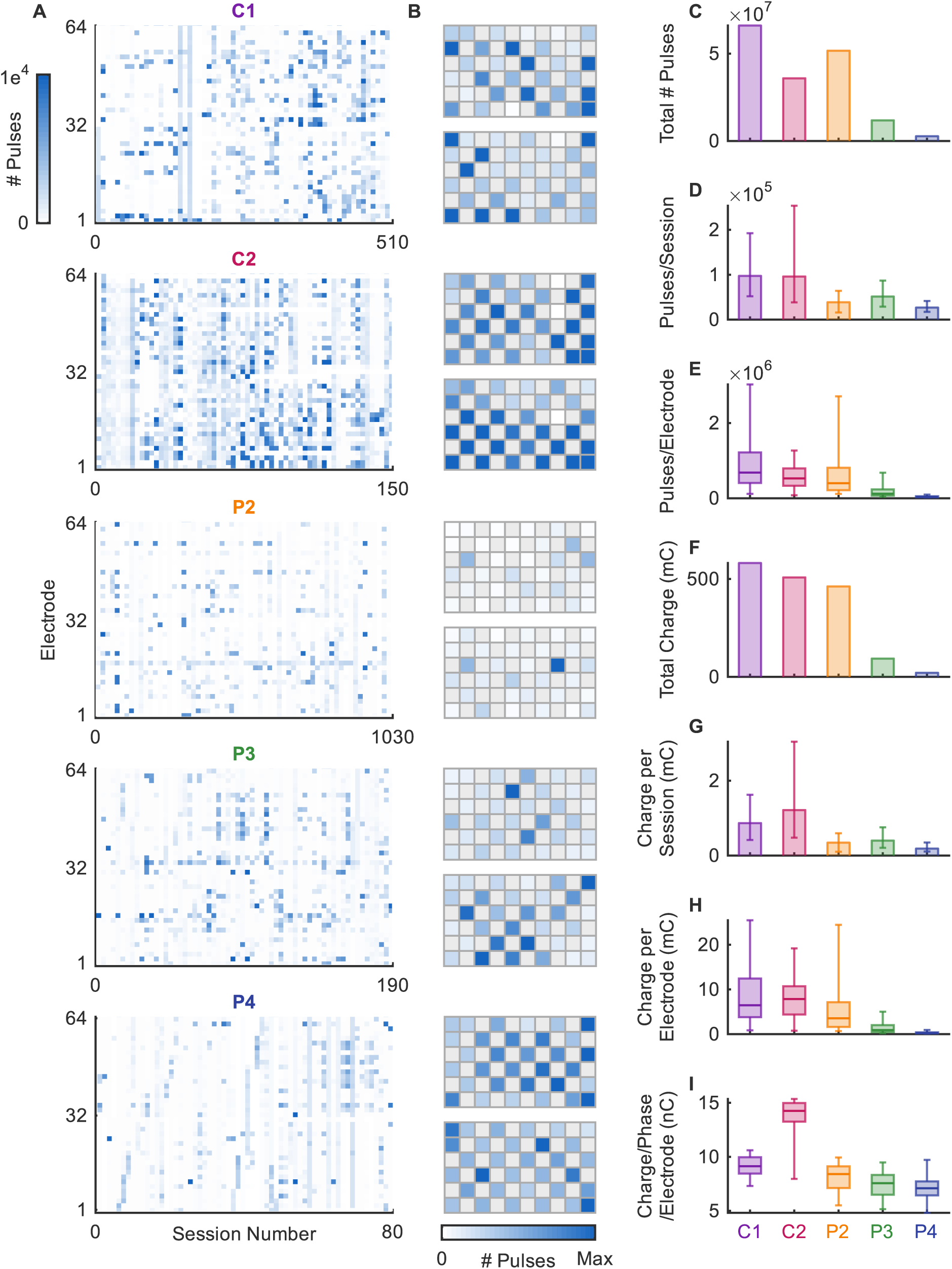
Detailed ICMS history for each participant. **A|** Heatmap of the number of pulses delivered per electrode (row) and per session (column) for each participant where the color of each cell corresponds to the number of pulses given. Color is capped at 1000 pulses per session for visualization purposes. **B|** Number of pulses on each electrode normalized to the number of pulses on the electrode receiving the most stimulus pulses. Disconnected electrodes are shown in gray. **C-I|** Summary box plots for total number of pulses, pulses per session, pulses per electrode, total charge, charge per session, charge per electrode, and charge per phase per electrode shown for each participant. Box and whisker indicate 5^th^, 25^th^, 50^th^, 75^th^, and 95^th^ percentiles.

**Supplementary Table 1:** Tactile thresholds before implantation and at the time of writing.

| Characteristic | C1 | C2 | P2 | P3 | P4 |
| --- | --- | --- | --- | --- | --- |
| **Threshold at implant (g)** |  |  |  |  |  |
| Thumb | 0.6 | >60 | 1.4 | 0.07 | >180 |
| Index | 0.6 | >60 | 10 | 0.07 | >180 |
| Middle | 2 | 60 | >180 | 6 | >180 |
| Ring | 1 | 26 | >180 | >300 | >180 |
| Pinky | 1 | 15 | >180 | 180 | >180 |
| Palm | 2 | - | 0.16 | 0.07 |  |
| **Threshold at time of writing (g)** |  |  |  |  |  |
| Thumb | 1 | >60 | 1.4 | - | >180 |
| Index | 0.6 | 26 | 10 | - | >180 |
| Middle | 0.6 | 15 | >180 | - | >180 |
| Ring | 0.6 | 26 | >180 | - | >180 |
| Pinky | 1 | 10 | >180 | - | >180 |
| Palm | 1 | >60 | 0.16 | - | >180 |
| **Change in threshold (g)** |  |  |  |  |  |
| Thumb | -0.4 | - | 0 | - | 0 |
| Index | 0 | - | 0 | - | 0 |
| Middle | 1.4 | 45 | 0 | - | 0 |
| Ring | 0.4 | 0 | 0 | - | 0 |
| Pinky | 0 | 5 | 0 | - | 0 |
| Palm | 1 | - | 0 | - | 0 |

**Supplementary Figure 2.**
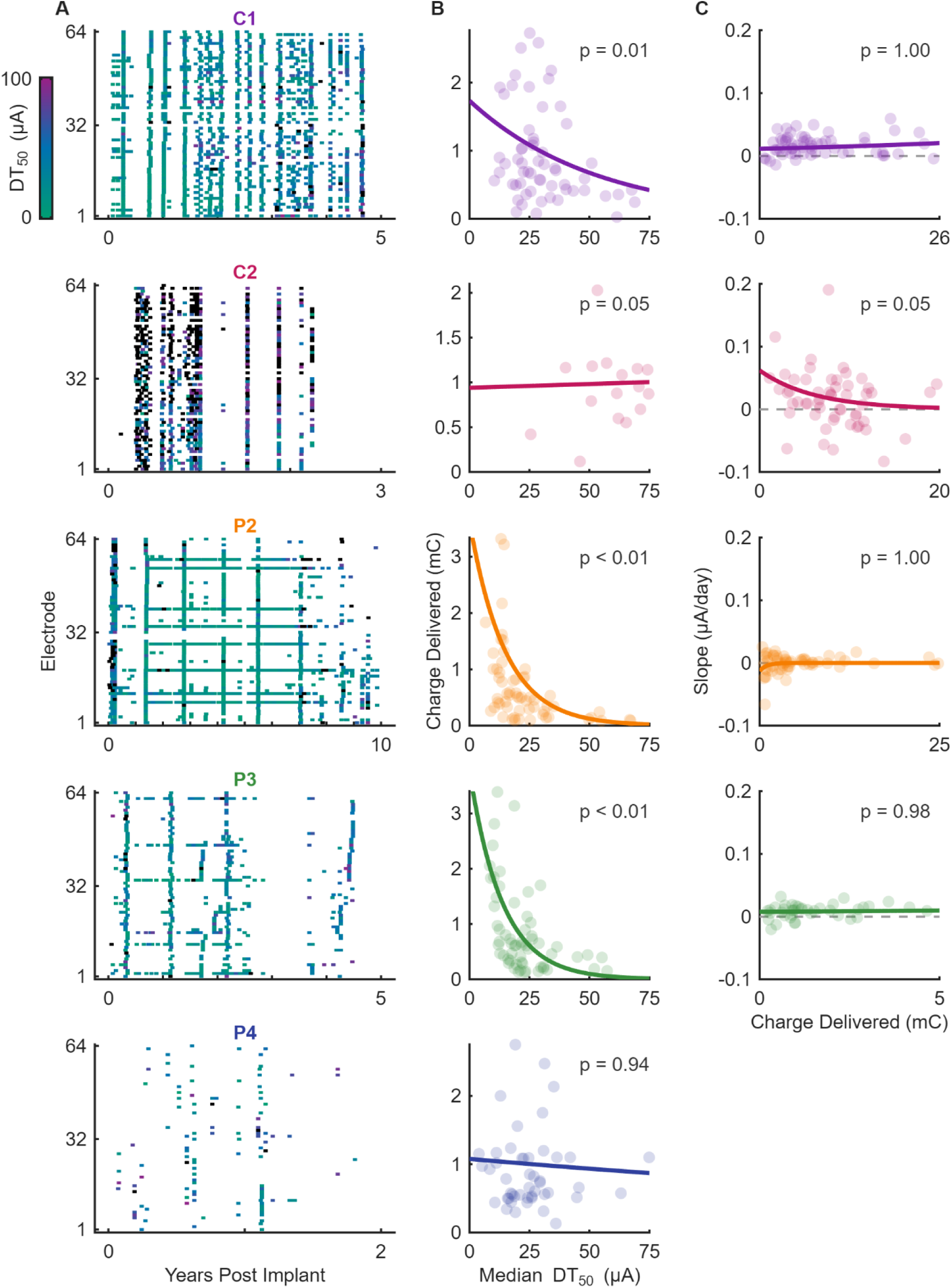
Detailed detection threshold history for each participant. **A|** Raster plots of detection thresholds for each electrode and participant. X-position of each tick indicates the time of the test, y-position indicates the electrode tested, and color indicates the estimated detection threshold corresponding to the color bar. Estimates that exceeded 100 μA are shown in black. **B|** Relationship between the median detection threshold for each electrode and the amount of charge delivered over the course of the study. **C|** Relationship between the total charge delivered and the change in detection threshold over time for each electrode. There were not enough detection thresholds collected from P4 to perform the regression analysis required to compute the slope for each electrode. P-values in B and C indicate Spearman correlations with Holm-Bonferroni post-hoc correction. Note that 2^nd^ order polynomials were fit to data for all participants in **B** and **C**.

**Supplementary Figure 3.**
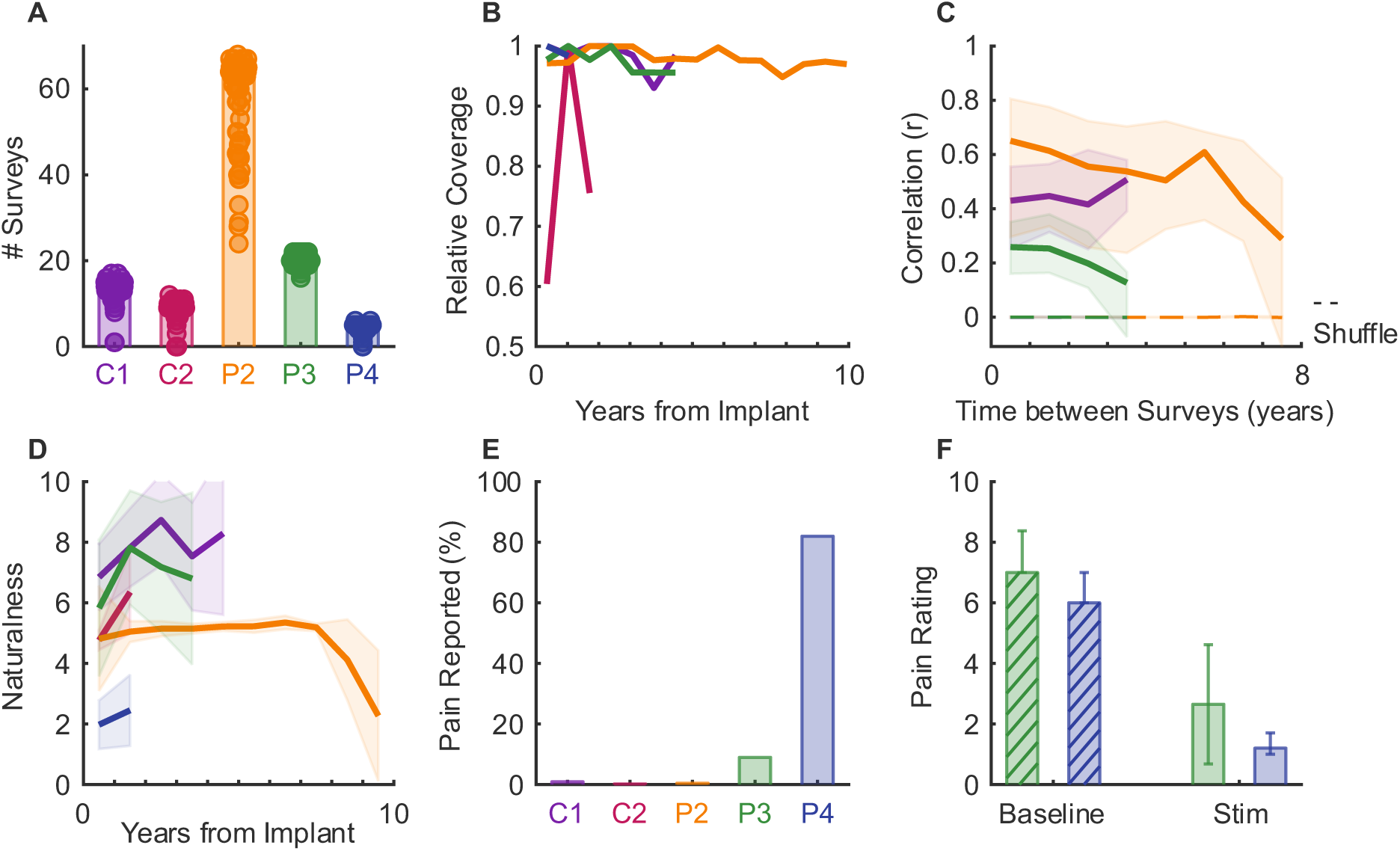
Perceptual stability. **A|** Number of surveys performed for each electrode and participant. **B|** Relative spatial coverage of the hand for each participant over time normalized to the maximum coverage reported. **C|** Similarity of quality reports within each electrode across multiple surveys organized by time between surveys. Shuffled controls are shown for each participant with dashed lines. Line indicates median and shaded area indicates the inter-quartile range. No data shown for participants C2 and P4 since there was an insufficient number of surveys. **D|** Mean (line) and standard deviation (shaded area) of the reported naturalness (0 = artificial, 10 = natural) across all electrodes over time for each participant. **E|** Frequency of a reported pain sensation in response to stimulation. **F|** Pain rating (0 = no pain, 10 = worst possible pain) for spontaneous pain reported at the start of a test session (baseline) and in response to single-electrode stimulation (stim-related) for participants P3 and P4.

**Supplementary Figure 4.**
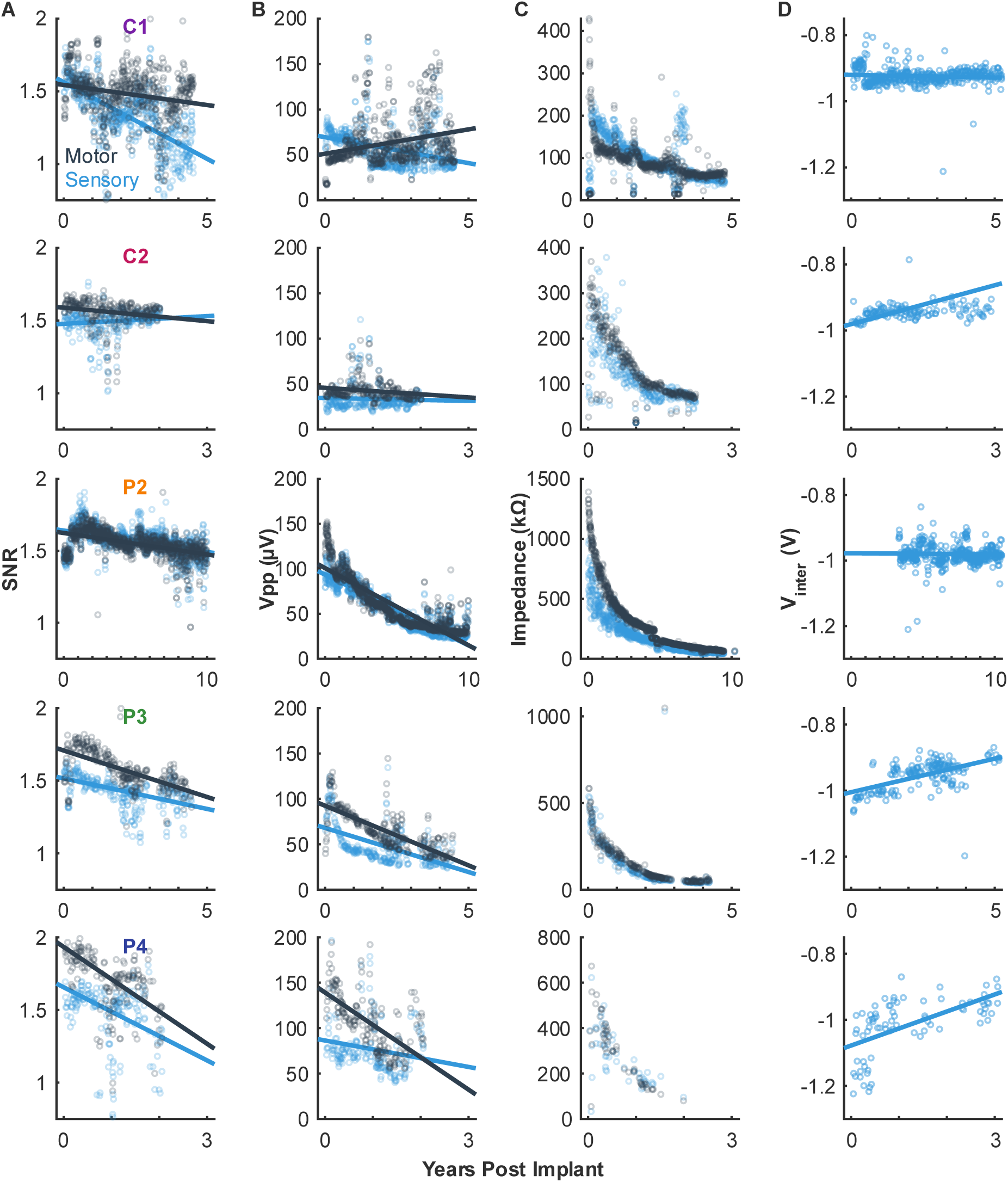
Electrode health and functionality. Median values across all electrodes on the sensory and motor arrays, respectively. **A|** Signal to noise ratio, **B|** peak to peak voltage, **C|** 1 kHz impedance, and **D|** interphase voltage over time for each participant with lines of best fit. For all plots, values are grouped by motor or sensory and the median value on a given day shown as individual points with the line of best fit shown over time.

**Supplementary Figure 5.**
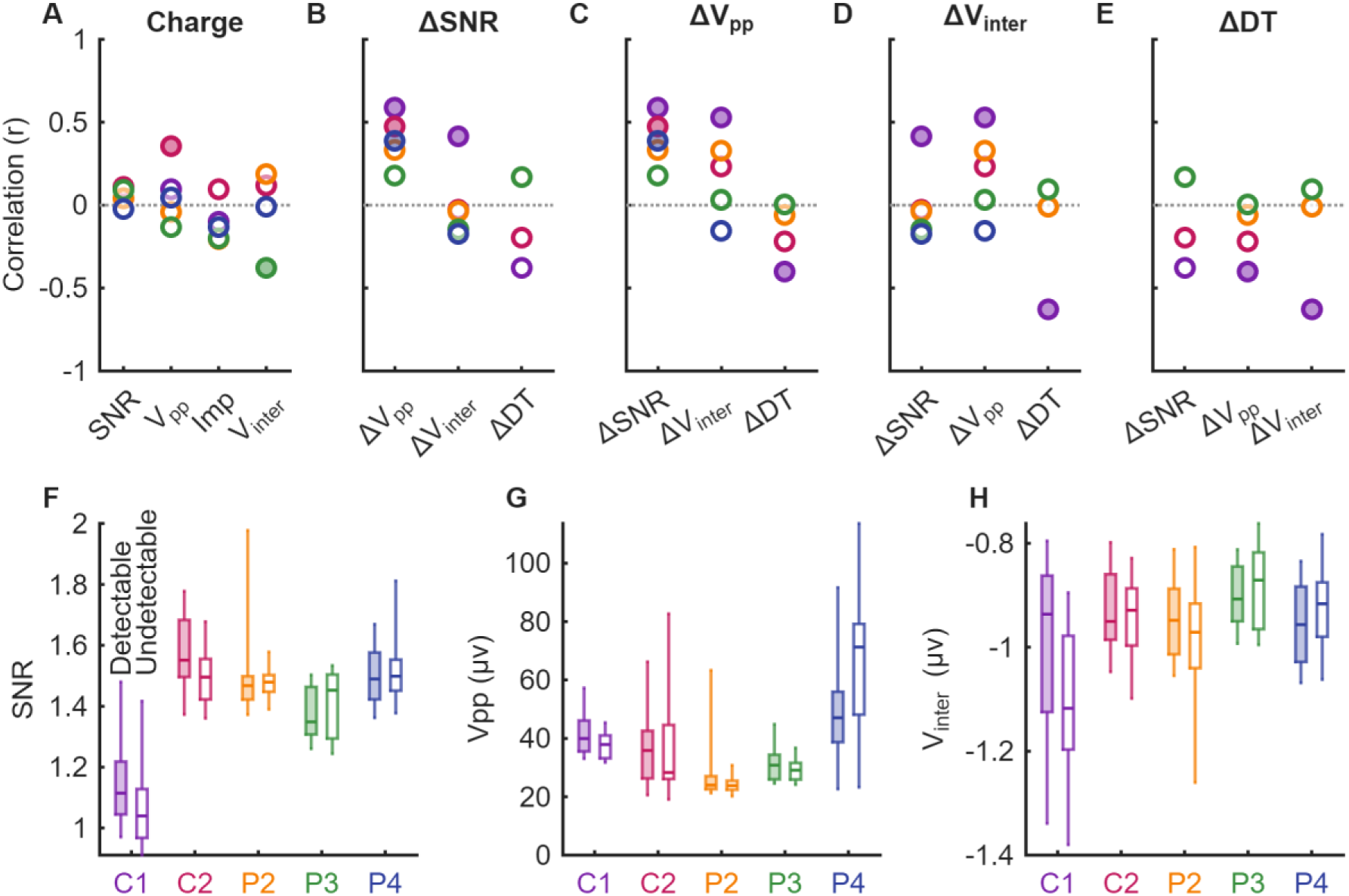
Relationships between changes in electrode health metrics. **A|** Correlations between the amount of charge delivered to each electrode and signal quality metrics. Filled circles indicate statistical significance. **B-E|** Correlation between ΔSNR, ΔV_pp_, Δinterphase voltage, and Δdetection threshold compared against all other metrics respectively. **F-H|** SNR, Vpp, and interphase voltage for each participant over the most recent 250 days split by whether electrodes were detectable or not. 2-way ANOVA results: **E**: Detectable[1], F=2.58, p=0.109; Participant[4], F=93.37, p<0.01. **F**: Detectable[1], F=1.0, p=0.317; Participant[4], F=37.59, p<0.01. **G**: Detectable[1], F=1.25, p=0.265; Participant[4], F=9.25, p<0.01.

**Supplementary Figure 6.**
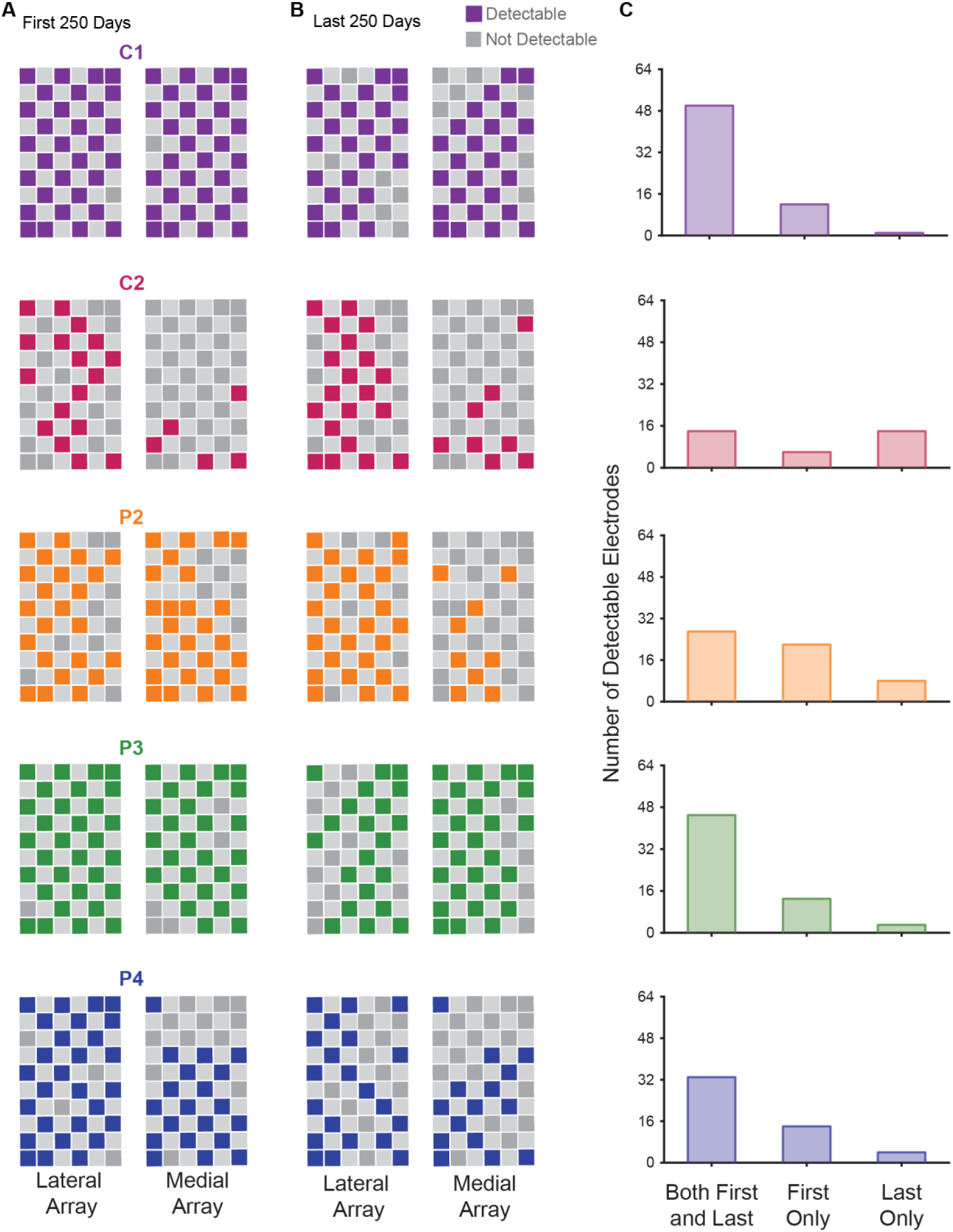
Relationships between array location and electrode detectability. Colored squares indicate the array location of detectable electrodes, calculated by the median detection threshold from the **A|** first 250 days to **B|** the most recent 250 days of data collection for each participant.

## Notes

### Clinical Trial

NCT01894802

### Author Declarations

This clinical trial includes participants at both the University of Pittsburgh and the University of Chicago. The study is conducted under an investigational device exemption from the US Food and Drug Administration, institutional review board protocols at both sites, and details of the trial are listed on clinicaltrials.gov (NCT01894802). Safety data and scientific progress were presented bi-annually to an independent data and safety monitoring board composed of experts in the field.

